# Methods for considering equality and equity implications in horizon scanning for medicines and healthcare innovations: a scoping review

**DOI:** 10.1101/2024.10.28.24316274

**Authors:** Chizoba Oparah, Tafadzwa Patience Kunonga, Claire Eastaugh, Gill Norman

**Author notes:** Corresponding author: Chizoba Oparah, NIHR Innovation Observatory, Population Health Sciences Institute, The Catalyst, Room 3.12, 3 Science Square Newcastle Helix Newcastle Upon Tyne, NE4 5TG.

## Abstract

**Background:** There is an increasing focus on inequity in healthcare and health outcomes. Early awareness of potential sources of inequity in access to and outcomes from innovative health technologies can support system preparedness and allow implementation of mitigations.

They may also be used to improve research inclusion.

**Objective:** To explore methods used to integrate equality and equity into horizon scanning for healthcare innovations, focusing on acceptability, polypharmacy, and multiple long-term conditions (MLTC).

**Design:** A scoping review followed Joanna Briggs Institute (JBI) guidelines to identify relevant methodologies for integrating equity into horizon scanning.

**Data sources:** Searches were conducted in MEDLINE, Embase, ProQuest, and WHO Global Index Medicus up to May 24, 2024.

**Eligibility criteria:** Studies were eligible if they presented methodologies for integrating equity and equality considerations into horizon scanning in health and care. Primary outcomes related to equity or equality, and secondary outcomes addressed acceptability, polypharmacy, and MLTC.

**Data extraction and synthesis:** Data were extracted on study characteristics, equity frameworks, and the integration of equity-related factors, including socioeconomic status, gender, and geographic location. A narrative synthesis was used to present the findings.

**Results:** Out of 951 records screened, three studies were included. The studies used varied horizon scanning methods, including scenario-building and foresight methodologies, and spanned multiple healthcare contexts such as precision oncology and complex paediatric care. Each study incorporated equity/equality by addressing the impact of emerging innovations on clinically vulnerable populations. Acceptability was found to be crucial for equitable implementation, particularly in precision oncology. However, managing complex health needs, especially in disadvantaged groups, is complicated by significant challenges such as polypharmacy and the presence of multiple long-term conditions.

**Conclusions:** Limited evidence highlighted a lack of consistent approaches to integrating equity into horizon scanning. While methods such as stakeholder engagement and scenario analysis showed promise, further research is needed to refine frameworks that better detect early indicators of inequity in healthcare innovation.

**Article summary:** *Strengths and limitations of this study:* Strengths

- Followed a robust and transparent methodology using the Joanna Briggs Institute (JBI) guidelines for scoping reviews.
- Comprehensive search strategy developed in collaboration with an experienced information specialist, covering multiple databases without restrictions.
- Dual independent screening and data extraction enhanced the reliability and consistency of the review process. Limitations

- Limited number of included studies and heterogeneity in methodologies and healthcare settings reduced the generalisability of findings.
- No critical appraisal of the quality of included studies, as the review focused on identifying methodologies rather than assessing study quality.

## Introduction

Horizon scanning is a systematic approach used in various sectors, including defence, environment, and healthcare, to identify emerging trends and innovations.[1] It involves the early detection and assessment of important developments, prioritizing resources and investments in innovation.[2] In healthcare, the need for horizon scanning is highlighted by the continued unmet medical need for new medicines, especially for conditions such as cancer, immunological diseases, and orphan diseases.[3] Additionally, the growing popularity of mobile health (mHealth) solutions and wearable devices necessitates the systematic identification and assessment of these emerging technologies for their sustained impact on healthcare.[4] While the primary goal of horizon scanning is to enhance preparedness and efficiency, it is essential to critically assess how it may be used to improve equality and equity in healthcare systems.

Equality and equity represent distinct approaches to addressing fairness and justice.[5] While equality emphasises uniform treatment and equal access to resources, opportunities, and rights for everyone regardless of individual circumstances, equity prioritises the distribution of resources and opportunities based on the specific needs and disadvantages of different groups.[5] Equality aims for sameness, treating everyone alike, while equity aims for fairness, ensuring that everyone has what they need to thrive, thus addressing systemic differences and levelling the playing field.[5] For instance, while equality might entail providing the same medical services to all, regardless of socioeconomic status, equity would involve allocating resources in a way that reduces health differences between different social groups, ensuring that those with fewer advantages receive the support necessary for better health outcomes.[5]

Existing literature has highlighted the importance of considering equality and equity in healthcare innovation processes to mitigate disparities in access, utilisation, and health outcomes.[6] However, there remains a gap in understanding how equality and equity considerations are integrated into horizon scanning activities, especially concerning new medicines, devices, diagnostics, and digital innovations (DDD). This scoping review aims to address this gap by exploring what methods have been used to consider implications for equity and associated domains of acceptability, polypharmacy, and the presence of multiple long-term conditions (MLTC) in horizon scanning.

Acceptability, polypharmacy, and the presence of MLTC are critical equity considerations, as they disproportionately affect clinically vulnerable populations. Acceptability plays a pivotal role in ensuring that healthcare innovations are perceived as appropriate, suitable, and accessible by diverse groups, thus enhancing the equitable uptake and delivery of healthcare services.[7] Polypharmacy, which refers to the concurrent use of multiple medications, can exacerbate health disparities, particularly in populations with lower health literacy, as it increases the risk of adverse drug reactions and medication non-adherence.[8] MLTC, defined as the coexistence of multiple health conditions in an individual, complicates the delivery of coordinated care, often contributing to unequal health outcomes, particularly in under-resourced settings.[9] These factors emphasise the need to integrate equity-focused methods in horizon scanning processes to ensure that healthcare innovations are both effective and accessible to all segments of the population.

By addressing this comprehensive research question, the scoping review aims to provide insights into the methods employed in horizon scanning to consider equality and/or equity implications and associated factors, contributing to the advancement of healthcare horizon scanning practices.

### Review question

What methods have been used to consider implications for equality and equity, and the associated domains of acceptability, polypharmacy, and presence of MLTC, within horizon scanning?

### Objectives

- To identify existing approaches for integrating equality and equity considerations into horizon scanning processes.
- To explore how acceptability, polypharmacy, and the presence of MLTC are addressed in the context of horizon scanning in relation to equality and equity.

### Eligibility criteria

Studies were included if they explored methodologies for incorporating equality and equity considerations within horizon scanning in health and care. Specifically, studies that presented frameworks or methods for integrating these considerations into horizon scanning processes were selected, while those not focused on these areas were excluded. Eligible studies had to include data related to horizon scanning and its integration with equality and equity. Primary measures were those addressing equality or equity, while secondary measures included assessments of acceptability, polypharmacy, and MLTC within horizon scanning. Full details of the inclusion criteria are available in Table 1.

**Table 1:** Eligibility Criteria.

| **Criterion** | **Inclusion** | **Exclusion** |
| --- | --- | --- |
| Types of studies | Studies presenting methods/frameworks for integrating equality and equity into horizon scanning processes in the broad field of health or care.<br>Studies published in English.<br>No country-based restrictions | Studies not focused on horizon scanning or not focused on equality/equity integration.<br>Studies in fields other than health or care.<br>Studies not published in English. |
| Types of data | Studies that include data related to horizon scanning processes and their integration with equality and equity. | Studies that lack relevant data on horizon scanning or equity considerations. |
| Types of measures | Primary: Types of measures that address equality or equity e.g., using PROGRESS-Plus framework.<br>Secondary: Studies that employ measures to assess acceptability, polypharmacy, and comorbidity domains within horizon scanning. | Studies that lack measures related to equality/equity, acceptability, polypharmacy, and managing multiple long-term conditions in horizon scanning. |
| Types of outcomes | Primary: Best practices and processes for addressing health equity or equality in horizon scanning using the PROGRESS- Plus framework.<br>Secondary: acceptability, polypharmacy, and multiple long term conditions considerations within horizon scanning methodologies. |  |

## Methods

This scoping review was conducted following the methodological guidance outlined by the Joanna Briggs Institute (JBI) for scoping reviews, [10] Critical appraisal was not conducted, as this research analysed how equality and equity considerations were integrated into horizon scanning, rather than the quality of framework implementation. The scoping review was reported according to the Preferred Reporting Items for Systematic Reviews and Meta-Analyses Extension for Scoping Reviews (PRISMA-ScR) guidelines,[11] and the protocol was registered with the Open Science Framework.[12]

### Search Strategy

The search strategy was designed in collaboration with an experienced information specialist (CE). The search was developed in MEDLINE (OVID). Elements of the equity grouping were taken directly from a paper by Hosking et al.,[13] which was located via the ISSG Search Filter Resource website.[14] This was then combined with additional relevant equity MeSH and key terms. The equity grouping was then combined with keyword terms for ‘horizon scanning’ and ‘foresight’. The final search strategy was peer-reviewed before being translated into Embase (OVID), ProQuest using the Social Science Premium Collection, and WHO’s Global Index Medicus. The searches were carried out from inception to 24 May 2024. No restrictions or limitations were imposed on the search. The search results were then combined and de-duplicated in EndNote. The full search strategies can be found in the Appendices 1 to 5.

### Study selection

Upon completing the systematic search, all identified citations were collated and duplicates removed using Endnote 21’s duplicate detection feature.[15] Two reviewers (CO, TPK) screened the title, abstract and full text levels was conducted using Rayyan.[16] Prior to formal screening, a pilot test was performed by randomly selecting 10% of the articles to ensure consistency and clarity in the application of inclusion criteria among the two reviewers. Any conflicts arising during the screening stages were discussed until consensus was reached. When necessary, a third opinion from another author (GN) was sought to achieve consensus. A PRISMA flow diagram was used to document the number of studies identified, screened, included, and excluded, with reasons for exclusion at the full-text stage provided.

### Data Extraction

Following the selection of studies, two reviewers (CO, TPK) piloted the data extraction form using one of the included studies to ensure the extraction of relevant information, and necessary modifications were made. Subsequently, the reviewers (CO, TPK) independently extracted the data on the remaining included studies and compiled the agreed extraction using Microsoft Excel. The extracted data included specific details of study characteristics (author, year, country), type of study, primary and secondary objectives, stages of horizon scanning, key technologies or interventions evaluated, equity and equality considerations, and any recommendations or implications for practice and policy. The scoping review adopted the PROGRESS-Plus framework to guide the exploration of equality and equity implications within horizon scanning processes.[17] PROGRESS plus factors include, Place of residence, Race/ethnicity/culture, Occupation, Gender/sex, Religion, Education, Socioeconomic status, Social capital, Plus other characteristics (e.g., age, disability, sexual orientation).[17]

### Data Analysis and presentation

The extracted data were synthesised using a narrative approach. The synthesis involved identifying patterns and themes across the studies regarding how equity and equality considerations were integrated into horizon scanning processes. Key themes, including the treatment of acceptability, polypharmacy, and MLTC, were identified, and mapped to the methods employed in horizon scanning. In addition, the synthesis examined how the studies operationalised the PROGRESS-Plus factors to address broader social determinants of health and ensure equity/inequity in the assessment of healthcare innovations.

## Results

### Study selection

A systematic search across electronic databases identified 1,152 records. After the removal of 201 duplicates, 951 unique records remained and were advanced to the initial screening phase. Title and abstract screening led to the exclusion of 935 studies. Consequently, 16 studies were retained for a more detailed full-text evaluation. Thirteen studies were excluded with reasons at this stage (see Appendix 6). Ultimately, 3 studies,[18–20] were included in the review (Fig.1).

**Fig 1.**
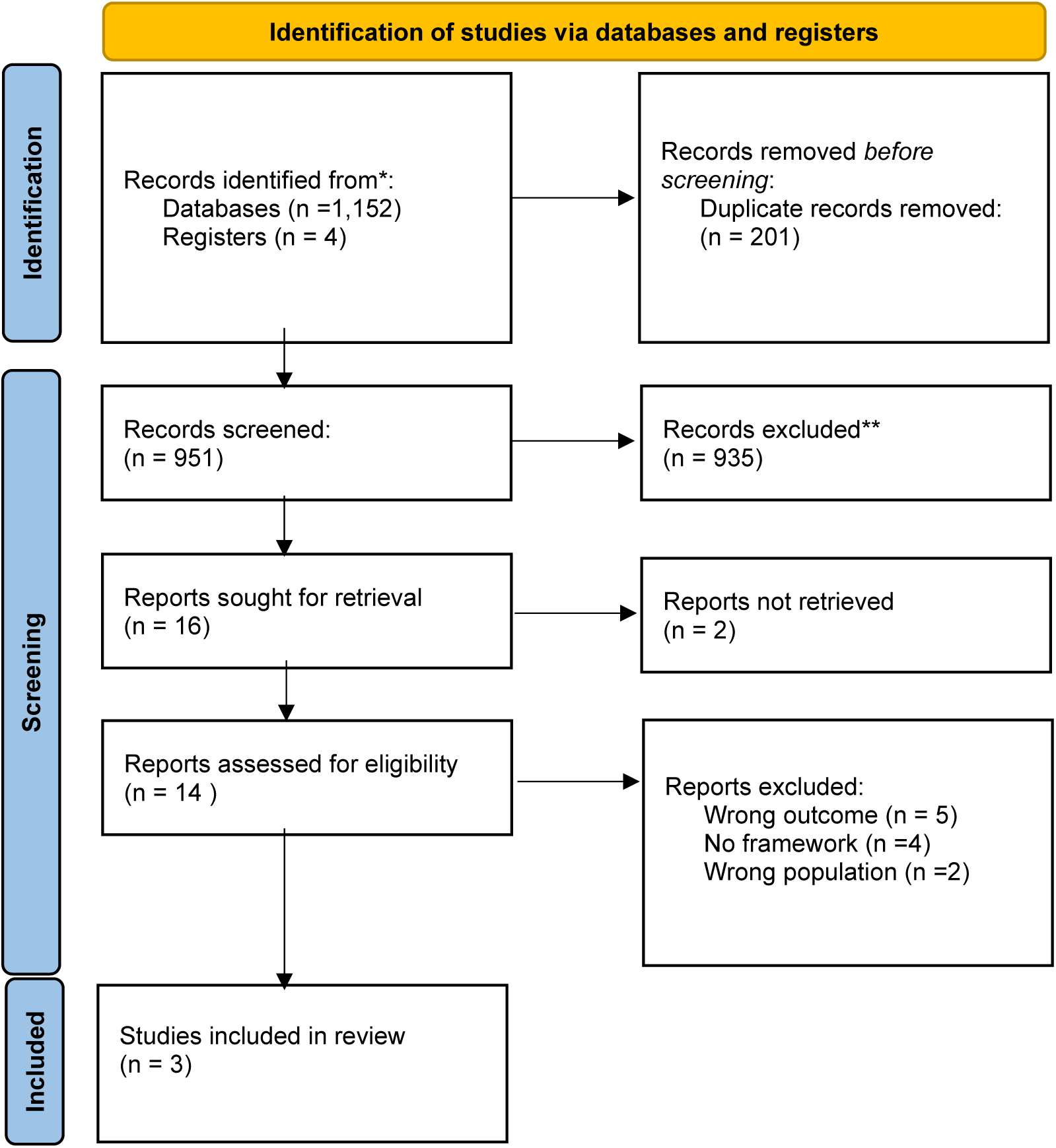
Flow diagram of included studies.

### Characteristics of included studies

The included studies, published between 2019 and 2024, span multiple countries and offer heterogenous insights into healthcare innovations. Studies were conducted in multiple European countries as part of the EURO-HEALTHY project,[18] Belgium,[20] and Canada.[19] The study designs varied, with one study,[18] employing a socio-technical scenario-building approach, another,[20] using a foresight methodology, and the third

study,[19] conducting a horizon scan. These studies were situated in different healthcare contexts: The study by Alvarenga and colleagues,[18] focused on the broader European public health landscape, Schmitt and colleagues,[20] focused on the integration of precision oncology into routine cancer care in Belgium, and Jones and colleagues,[19] on the care of children and youth with medical complexity in Canada. The healthcare innovations discussed include the integration of medical technologies and innovations to address health inequalities,[18] precision medicine in oncology,[20] and emerging models of care, communication systems, and genetic testing for medically complex paediatric populations.[19] Descriptive information on each individual study is presented in Table 2.

**Table 2:** Characteristics of included studies.

| Study ID | Study Characteristics |  | Study Context |  |  |
| --- | --- | --- | --- | --- | --- |
|  | Country | Type of study | Objectives | Setting | Emerging Technology Discussed |
| Alvarenga et al., 2019[18] | Various European countries (part of the EURO-HEALTHY project) | Socio-technical scenario-building approach | To develop future scenarios for population health inequalities in Europe | Broader European public health landscape | Integration of medical technologies to address health inequalities |
| Jones et al., 2024[19] | Canada | Horizon scanning | To identify and prioritise emerging technologies affecting care for children and youth with medical complexity. | Canadian health system focusing on paediatric care | Communication systems, genetic testing, and models of care for children with medical complexity |
| Schmitt et al., 2024[20] | Belgium | Foresight study | To explore factors influencing the equitable implementation of precision medicine in routine cancer care | Belgian cancer care system | Precision oncology, personalised cancer care, AI-supported decision-making tools, genomic data sharing |

### Themes and Findings

#### Horizon Scanning Methods Used

The studies employed various horizon scanning methods, reflecting the diversity of contexts and goals. Detailed descriptions of these methods are provided in Table 3. Alvarenga and colleagues,[18] used a three-stage socio-technical approach involving a Web-Delphi process, expert workshops, and scenario building to develop future scenarios for population health inequalities in Europe. This method allowed for a comprehensive and participatory exploration of potential health inequalities. Schmitt and colleagues,[20] employed a foresight methodology using the DESTEP framework (Demographic, Economic, Societal, Technological, Environmental, and Political/Policy factors) to assess the factors influencing the equitable implementation of precision medicine in Belgium. This approach involved a systematic literature review, expert surveys, and workshops to anticipate and address barriers to equitable healthcare. Jones and colleagues,[19] conducted a horizon scan as part of the Canadian Agency for Drugs and Technologies in Health (CADTH) Watch List, using a modified James Lind Alliance priority-setting approach to identify and prioritize emerging technologies and issues affecting the care of children and youth with medical complexity in Canada. This method was grounded in stakeholder engagement, ensuring that the scan reflected diverse perspectives.

**Table 3:** Horizon Scanning and Foresight Methods.

| Study ID | Horizon Scanning and Foresight Methods |  |  |  |
| --- | --- | --- | --- | --- |
|  | Horizon-Scanning Method Used (e.g., literature review, expert panels, Delphi method) | Foresight Method Used (e.g., scenario planning, trend analysis, visioning) | Description of the Methodology (detailed description of how the methods were applied) | Stakeholders Involved (e.g., policymakers, scientists, ethicists, public representatives) |
| Alvarenga et al., 2019[18] | Web-Delphi process, expert workshops | Scenario planning | A three-stage socio-technical approach involving a Web-Delphi process and expert workshops to develop future scenarios for population health inequalities across Europe. | Policymakers, scientists, and public health experts |
| Jones et al., 2024[19] | Priority-setting approach | Horizon scanning | Horizon scan conducted using a modified James Lind Alliance priority-setting approach to identify and prioritize emerging technologies affecting care for children and youth with medical complexity. | Healthcare providers, caregivers, policymakers |
| Schmitt et al., 2024[20] | Literature review, expert survey | Foresight study<br>Demographic, Economic, Societal, Technological, Environmental, and Political/Policy factors (DESTEP) framework | A literature review and expert survey assessed contextual factors using the DESTEP framework to explore the equitable implementation of precision oncology in Belgium. | Cancer care experts, policymakers, researchers |

### Approaches for Integrating Equality and Equity Considerations into Horizon Scanning Processes

All three studies placed significant emphasis on integrating equity considerations into their respective horizon scanning processes, with specific details presented in Table 4. Alvarenga and colleagues,[18] incorporated equity by ensuring that the scenarios developed addressed how different political, economic, and social factors could influence health inequalities across Europe. The study highlighted the potential impact of these factors on various population groups, emphasising the need for policies that reduce disparities. Schmitt and colleagues,[20] focused on equity by assessing the contextual factors that could either facilitate or hinder the equitable implementation of precision medicine in Belgium. The study identified key economic and technological factors, such as dedicated healthcare budgets and data standardisation, as crucial for ensuring that precision oncology benefits are accessible to all segments of the population. Jones and colleagues,[19] addressed equity by prioritising technologies and issues that could reduce disparities in care for children and youth with medical complexity. The study emphasised the importance of equitable access to emerging healthcare innovations, particularly for populations in rural or underserved areas.

**Table 4:**
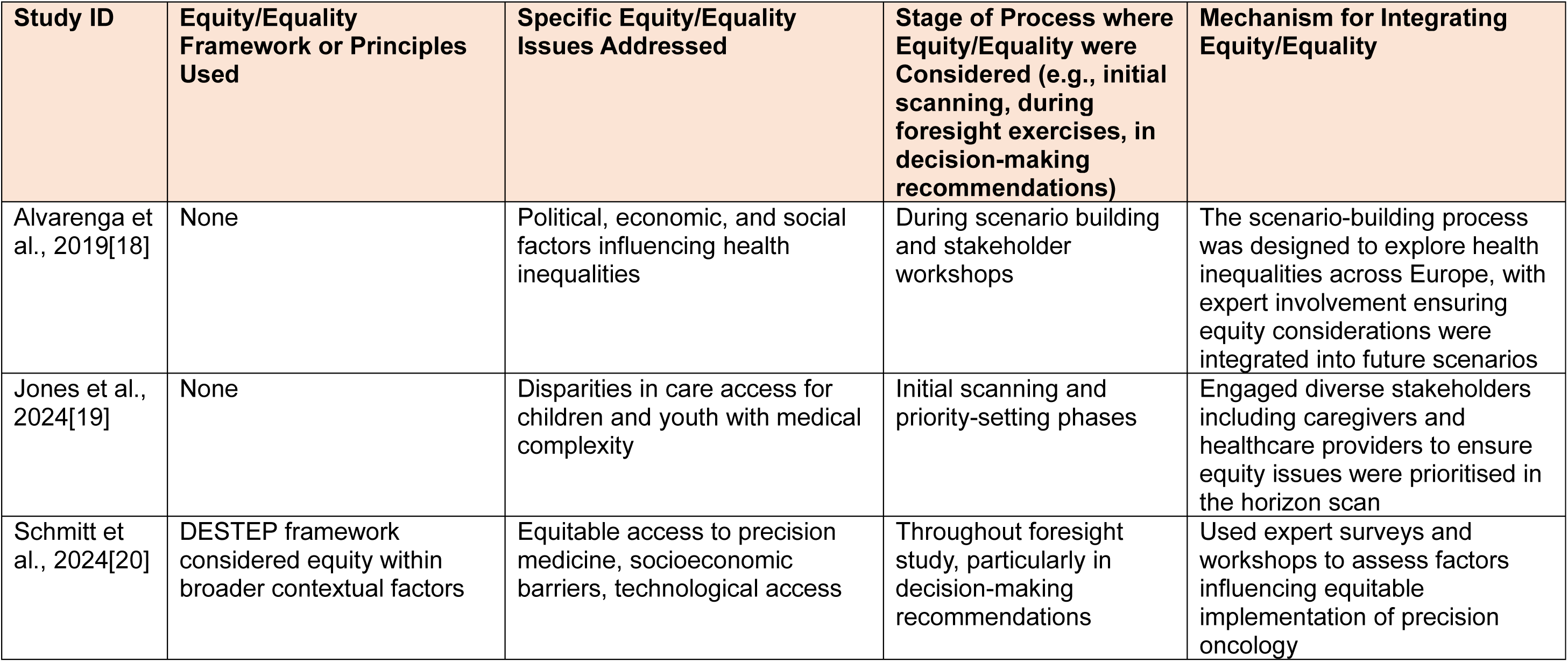
Equity considerations.

### Acceptability, Polypharmacy, and MLTC

Acceptability, polypharmacy, and MLTC were variably addressed across the studies. Acceptability was a critical consideration in all three studies; detailed information is available in Table 5. Alvarenga and colleagues,[18] indirectly addressed acceptability through the exploration of public health policies’ success, influenced by their acceptance among different stakeholders. Schmitt and colleagues,[20] addressed acceptability in the context of precision oncology, emphasising the need for new technologies to be accepted by both patients and healthcare providers to ensure successful implementation. Jones and colleagues,[19] considered acceptability by focusing on the challenges faced by caregivers in adopting new healthcare technologies and ensuring that these innovations align with the needs and capacities of families.

**Table 5:** Acceptability, Polypharmacy, and Multiple Long-Term Conditions.

| Study ID | Acceptability | Polypharmacy | Multiple Long-Term Conditions |
| --- | --- | --- | --- |
| Alvarenga et al., 2019[18] | Indirectly addressed by exploring how public health policies' success is influenced by their acceptance among different stakeholders, focusing on how future scenarios may impact diverse groups' views on health interventions. | Not specifically addressed. | Not specifically addressed. |
| Jones et al., 2024[19] | New healthcare technologies were evaluated for their alignment with the needs and capacities of families. | Polypharmacy was relevant in the context of managing multiple medications for children with medical complexity, with a focus on technologies that support effective medication management. | The study emphasised the need for coordinated care models to address the complex health conditions present in children with medical complexity. |
| Schmitt et al., 2024[20] | Emphasised the need for new technologies to be accepted by both patients and healthcare providers to ensure successful implementation. | Precision medicine was discussed as a potential solution for reducing polypharmacy risks by tailoring treatments to individual genetic profiles, thereby minimising unnecessary drug use. | Study highlighted as an important factor in managing multiple health conditions alongside cancer, with precision medicine offering a more targeted approach to treatment. |

Polypharmacy was particularly relevant in the studies by Schmitt and colleagues,[20] and Jones and colleagues.[19] Schmitt and colleagues,[20] discussed how precision medicine could reduce the risks associated with polypharmacy by tailoring treatments to individual genetic profiles, thereby minimising unnecessary drug use. Jones and colleagues,[19] addressed polypharmacy in the context of managing complex medication regimens for children with multiple health conditions, highlighting the role of emerging technologies in supporting effective medication management.

The presence of MLTC was a central theme in the studies by Schmitt and colleagues,[20] and Jones and colleagues.[19] While Schmitt and colleagues,[20] explored how precision medicine could be integrated into the care of patients with multiple health conditions, ensuring that comorbidities are managed alongside cancer treatments, Jones and colleagues,[19] emphasised the need for coordinated care models that address the complex comorbidities often present in children with medical complexity, advocating for technologies and care strategies that support holistic and integrated care.

## Discussion

### Summary of Findings

Our review synthesised methods from three heterogenous studies, emphasising the importance of integrating equity/equality considerations in horizon scanning to efficiently address health disparities. The main methodological approaches common to all studies can be categorised into: Stakeholder engagement, Expert surveys, and Scenario analysis.

These were highlighted across all studies through the use focus groups, web-Delphi, priority-setting sessions, and workshops to identify and validate future-oriented evidence that explore and address complex health issues.[18–20] Overall, the included studies suggest that employing horizon scanning methods and foresight methodology approaches can help to identify potential key drivers affecting the gradual rise of healthcare inequalities/inequities.[5] These methods allow researchers and policymakers to evaluate the potential impacts of emerging technologies and healthcare practices on health inequalities/inequities using scenario-building to explore different structural contexts. These scenarios are often validated in expert workshop formats, reinforcing the importance of interdisciplinary collaboration and diverse stakeholder input. This process highlights the necessity of employing varied methodologies while integrating equity/equality considerations to address healthcare inequalities comprehensively.[20]

Despite the strengths of these methodologies, a significant gap remains in their ability to detect weak signals—subtle, early indicators of potential inequalities/inequities that may not yet be fully visible but could evolve into more significant drivers of disparities if not addressed.[21] These weak signals might include emerging trends in healthcare access, socio-economic shifts, or early-stage technological developments that disproportionately impact marginalised groups. They may also include signs of poor research inclusion of marginalised groups which could reduce the relevance, safety and effectiveness of novel technologies for people from these groups. Our review highlights the need for future horizon scanning methodologies to incorporate more sophisticated, equity/inequity-focused tools capable of capturing these early warning signals. This would enable a more proactive response to inequities/inequalities before they become entrenched, ensuring that healthcare innovations are accessible to all populations.[21]

### Comparisons with Existing Literature

Our findings align with the broader literature on health equity/equality, which highlights the critical importance of integrating equity and/or equality considerations into healthcare innovation processes. The use of frameworks such as DESTEP in the study by Schmitt and colleagues,[20] reflects a systematic approach to evaluating the socio-economic, technological, and environmental factors that influence health disparities. This aligns with existing research advocating for the inclusion of equity in health assessments to prevent the widening of health inequalities.[5 17] The consideration of acceptability, polypharmacy, and MLTC within horizon scanning processes is crucial, given the complexities associated with managing chronic conditions and multiple medications in diverse and vulnerable populations. The acceptability of healthcare innovations to diverse populations is crucial, as failure to account for cultural, social, and economic factors may result in reduced uptake of interventions among marginalized groups, ultimately undermining their effectiveness.[7] Polypharmacy is a key indicator of inequity of access, as minoritised and disadvantaged groups are disproportionately likely to experience polypharmacy due to systemic barriers in healthcare.[8] These barriers often lead to fragmented care, increasing the risk of adverse drug events and poorer health outcomes.[8] By considering polypharmacy as a proxy for inequity, horizon scanning can help identify innovations that improve access to appropriate care and reduce the burden of managing multiple medications among clinically vulnerable populations.[8] This approach is consistent with the literature on the challenges of polypharmacy and the importance of personalised, patient-centred care strategies.[8 9] Equally important is the consideration of MLTC, which often correlates with higher healthcare needs and complexities, particularly among socially and economically disadvantaged populations.[22]

### Implications for Practice and Policy

The findings from this review highlight the necessity of systematically incorporating equity and/or equality considerations into horizon scanning processes, utilising established frameworks such as PROGRESS-Plus,[17] or DESTEP,[20] to prevent the worsening of health inequalities/inequities. It is crucial for policymakers, researchers, and practitioners to engage diverse stakeholders, including patients and healthcare providers, to ensure that new technologies align with user needs and expectations, thereby enhancing their acceptability. Additionally, as the prevalence of chronic conditions and the associated challenges of polypharmacy and MLTC continue to rise, horizon scanning must rigorously evaluate how emerging technologies can effectively address these complex care needs, particularly in vulnerable populations and, conversely risks that these may be exacerbated by new technologies. This integrated approach will support the development of healthcare innovations that are not only effective but also equitably accessible across diverse groups.

### Strengths and Limitations

This scoping review has several methodological strengths. The review followed a systematic approach, adhering to JBI guidelines, which ensured that relevant studies were identified.

The use of focused search strategies, combined with duplicate reviewer screening and data extraction, enhanced the rigour and transparency of the review process. Furthermore, the integration of the PROGRESS-Plus framework allowed for a structured exploration of equity considerations across diverse healthcare contexts.

However, the review also presents limitations. The small number of included studies and the heterogeneity in their methodologies and healthcare settings pose challenges for the generalisability of the findings. This diversity limited our ability to synthesise the results into broader, more cohesive conclusions. Additionally, while the studies used different horizon scanning methodologies, the variability in their design and context makes it difficult to draw definitive insights on the effectiveness of specific approaches to integrating equity into horizon scanning.

### Future Research Directions

A gap identified in this review is the need for further empirical testing of horizon scanning methods to assess how well they can detect weak signals - subtle, early indicators of potential inequalities/inequities that may not yet be visible but could evolve into significant drivers of disparity.[21] Future research should aim to expand the evidence base on integrating equity and/or equality into horizon scanning by developing and validating more comprehensive frameworks tailored to this purpose. In addition, further investigation is needed into how horizon scanning processes can be adapted to better identify and address the challenges of polypharmacy and MLTC, especially in aging populations and those with complex health needs. Such research will be instrumental in refining horizon scanning methodologies to ensure that they support the development of equitable and effective healthcare systems globally.

## Conclusion

This scoping review emphasises the critical need for integrating equity/equality considerations into horizon scanning to ensure that healthcare innovations are both effective and accessible across diverse populations. Addressing the domains of acceptability, polypharmacy, and MLTC can enhance the equity/equality of healthcare systems, as these factors disproportionately affect vulnerable populations. However, current horizon scanning methods lack a robust framework for detecting early weak signals of emerging inequities, such as subtle shifts in healthcare access or socio-economic disparities. Developing and testing methods to identify these signals is crucial for proactively addressing health disparities.

While this review focused on key factors, other important equity/equality-related considerations, such as digital health disparities and the intersectionality of social determinants, remain underexplored.[23] Expanding horizon scanning to include these broader factors is essential for ensuring truly equitable healthcare innovations. Future research should aim to refine and broaden horizon scanning methodologies to better detect early inequities/inequalities and include a wider range of factors to comprehensively address health disparities. Such research should ideally be co-produced with patient and public collaborators, including representatives of groups who are under-represented in clinical research.

## Supporting information

PRISMA-ScR-Checklist_V0.1

## Article Information Contributors

CO codesigned the protocol, methodology, writing of the original draft, visualisation of results, the overall project administration and is the guarantor of this work. TPK codesigned the protocol, undertook all stages of the review and cowrote the paper. CE undertook the searches for the review, codesigned the protocol and cowrote the paper. GN was responsible for the conceptualisation of this project, contributed to the analysis of review data and cowrote the paper.

## Acknowledgements

We wish to thank Alex Inskip, Research Assistant Information Specialist, and Sheila Wallace, Research Fellow Information Specialist, at the NIHR Innovation Observatory for peer-reviewing the search strategy. We would also like to thank Jane Mcdermott from the University of Manchester who generously provided knowledge and expertise.

## Ethics

As this study involves the synthesis of existing literature, ethical approval is not required.

## Disclaimer

The views expressed are those of the author(s) and not necessarily those of the NIHR or the Department of Health and Social Care.

## Patient and public involvement

Patients and/or the public were not involved in the design, or conduct, or reporting, or dissemination plans of this research.

## Funding/support

This study/project is funded by the National Institute for Health and Care Research (NIHR) (NIHRIO/project reference HSRIC-2016-10009).

## Role of the Funder/Sponsor

The funding sources had no role in the design and conduct of the study; collection, management, analysis, and interpretation of the data; preparation, review, or approval of the manuscript; and the decision to submit the manuscript for publication.

## Competing interests

None declared.

## Data availability statement

All data relevant to the study are included in the article or uploaded as supplemental information.

## Appendices

## Appendix 1 Summary of results

Appendix 1: Summary of results
| Database | Date of search | # results |
| --- | --- | --- |
| MEDLINE | 24/05/24 | 163 |
| Embase | 24/05/24 | 339 |
| ProQuest | 24/05/24 | 576 |
| WHO's Global Index Medicus | 24/05/24 | 73 |
| **Total before deduplication** |  | **1,152** |
| **Total after deduplication** |  | **951** |

## Appendix 2 MEDLINE (OVID)

Database(s): **Ovid MEDLINE(R) and Epub Ahead of Print, In-Process, In-Data-Review & Other Non-Indexed Citations, Daily and Versions 1946 to May 23, 2024**

Search Strategy:

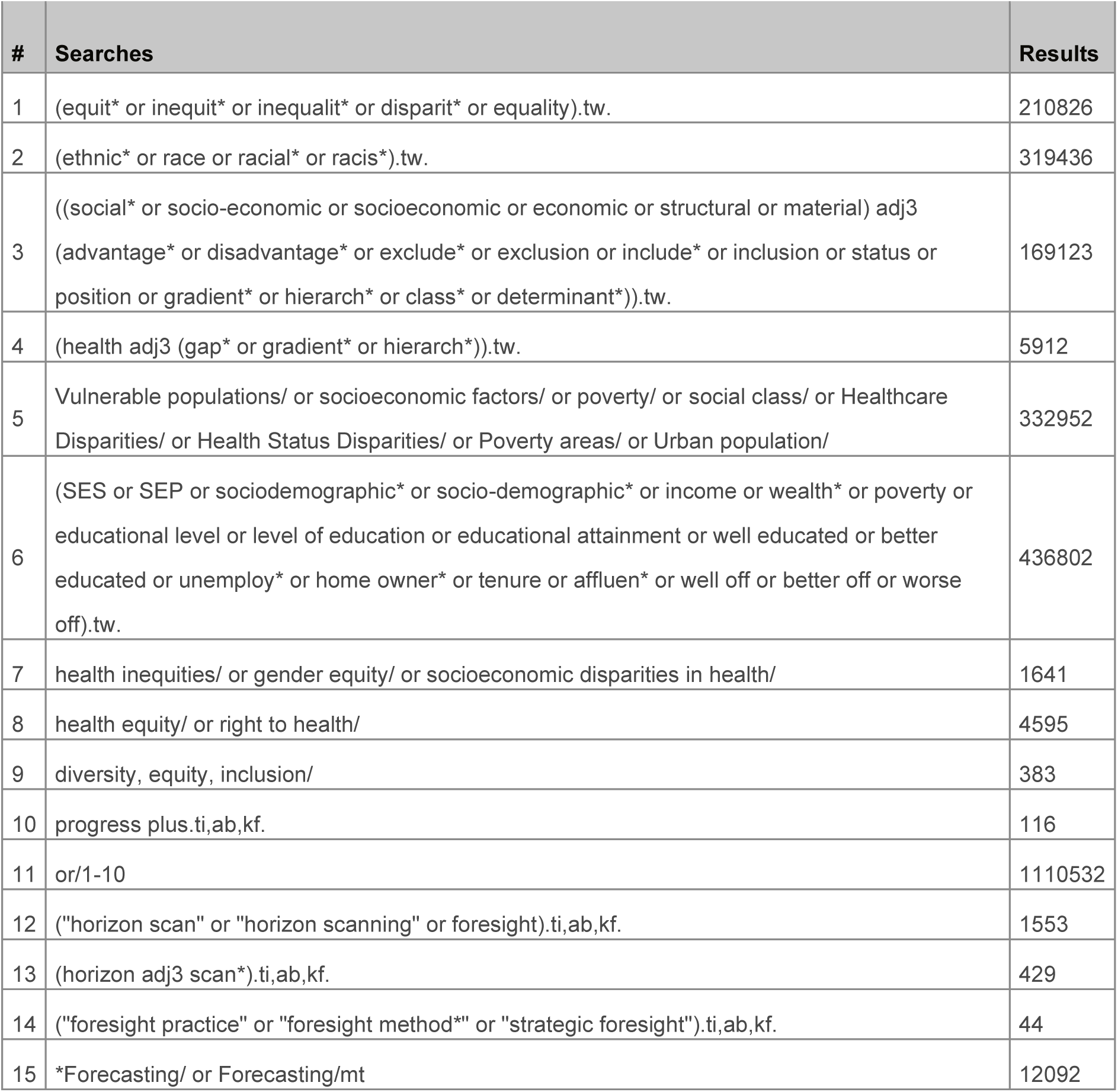
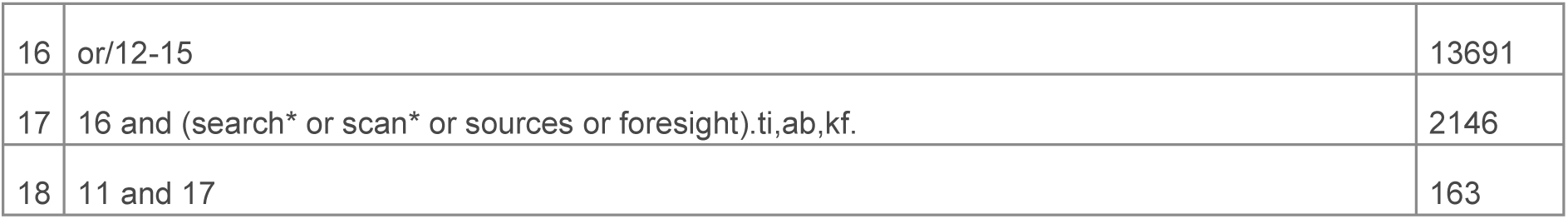

## Appendix 3 Embase (OVID)

Database(s): **Embase** 1974 to 2024 May 23

Search Strategy:

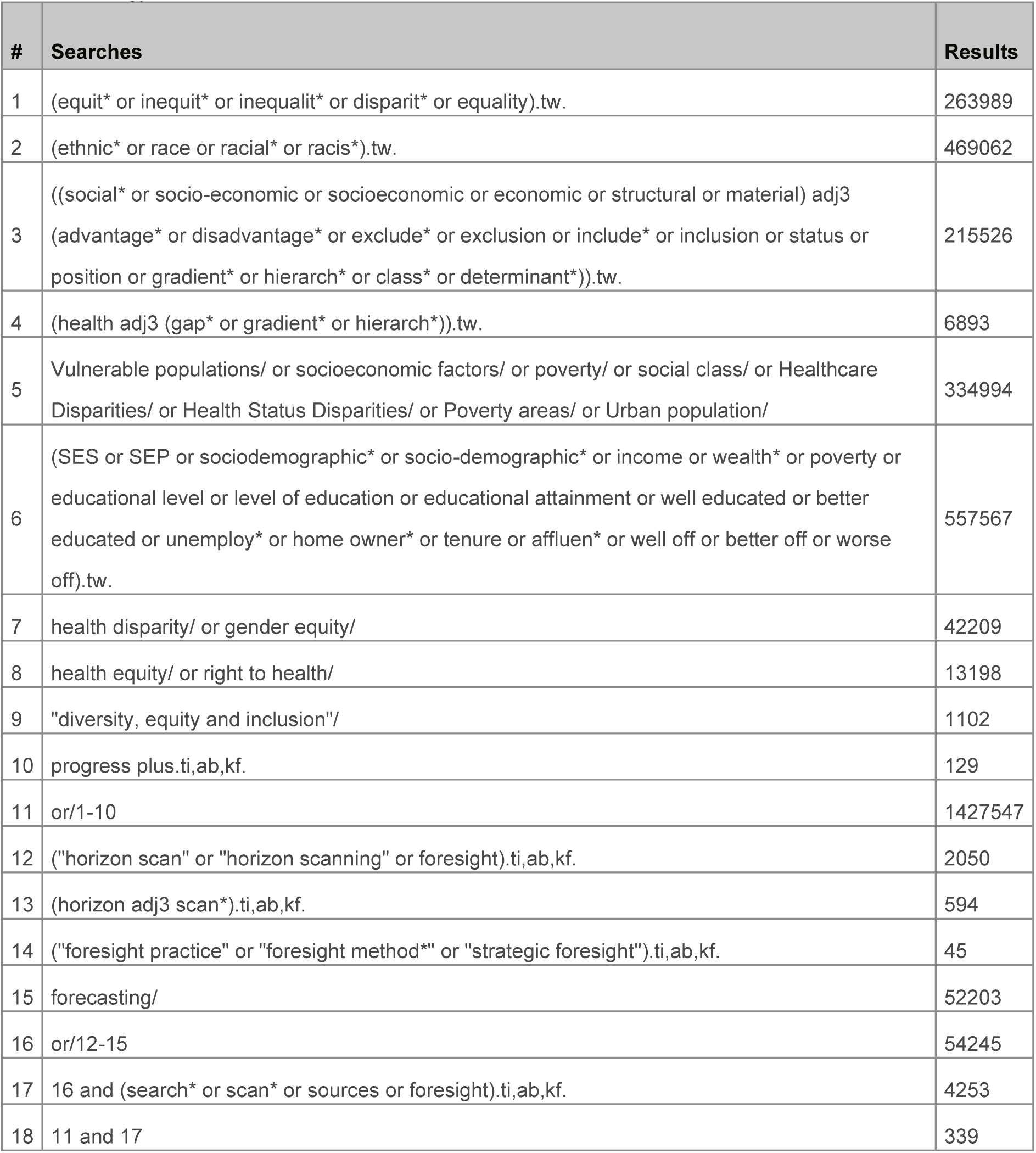

## Appendix 4 ProQuest

Social Science Premium Collection: Criminal Justice Database (1981 - current) | Education Collection (1966 - current) | International Bibliography of the Social Sciences (IBSS) (1951 - current) |Library & Information Science Collection (1969 - current) | Linguistics Collection (1973 - current) | Politics Collection (1909 - current) | Social Science Database | Sociology Collection (1952 - current)

Excluding due to platform export restrictions: National Criminal Justice Reference Service (NCJRS) Abstracts Database (1975 - current) | Criminology Collection (1975 - current)

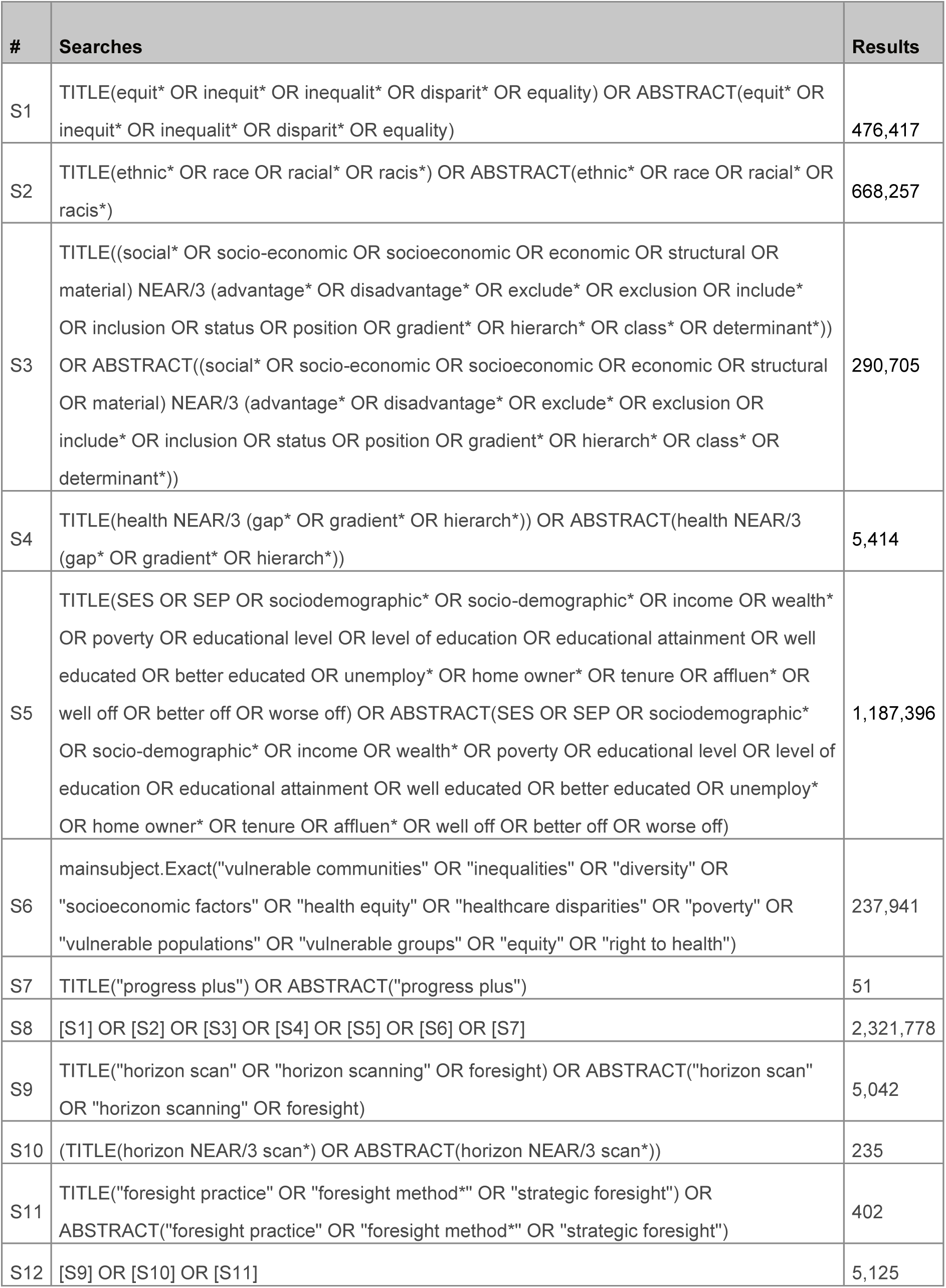
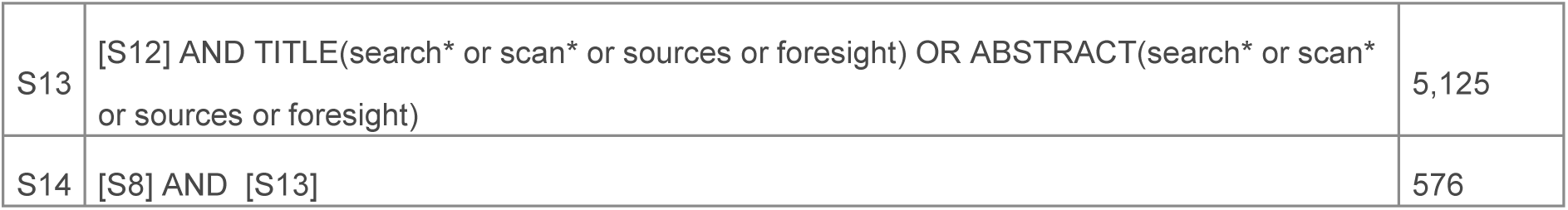

| # | Searches | Results |
| --- | --- | --- |
| S1 | TITLE(equit* OR inequit* OR inequalit* OR disparit* OR equality) OR ABSTRACT(equit* OR inequit* OR inequalit* OR disparit* OR equality) | 476,417 |
| S2 | TITLE(ethnic* OR race OR racial* OR racis*) OR ABSTRACT(ethnic* OR race OR racial* OR racis*) | 668,257 |
| S3 | TITLE((social* OR socio-economic OR socioeconomic OR economic OR structural OR material) NEAR/3 (advantage* OR disadvantage* OR exclude* OR exclusion OR include* OR inclusion OR status OR position OR gradient* OR hierarch* OR class* OR determinant*))<br>OR ABSTRACT((social* OR socio-economic OR socioeconomic OR economic OR structural OR material) NEAR/3 (advantage* OR disadvantage* OR exclude* OR exclusion OR include* OR inclusion OR status OR position OR gradient* OR hierarch* OR class* OR determinant*)) | 290,705 |
| S4 | TITLE(health NEAR/3 (gap* OR gradient* OR hierarch*)) OR ABSTRACT(health NEAR/3 (gap* OR gradient* OR hierarch*)) | 5,414 |
| S5 | TITLE(SES OR SEP OR sociodemographic* OR socio-demographic* OR income OR wealth* OR poverty OR educational level OR level of education OR educational attainment OR well educated OR better educated OR unemploy* OR home owner* OR tenure OR affluen* OR well off OR better off OR worse off) OR ABSTRACT(SES OR SEP OR sociodemographic* OR socio-demographic* OR income OR wealth* OR poverty OR educational level OR level of education OR educational attainment OR well educated OR better educated OR unemploy* OR home owner* OR tenure OR affluen* OR well off OR better off OR worse off) | 1,187,396 |
| S6 | mainsubject.Exact("vulnerable communities" OR "inequalities" OR "diversity" OR "socioeconomic factors" OR "health equity" OR "healthcare disparities" OR "poverty" OR "vulnerable populations" OR "vulnerable groups" OR "equity" OR "right to health") | 237,941 |
| S7 | TITLE("progress plus") OR ABSTRACT("progress plus") | 51 |
| S8 | [S1] OR [S2] OR [S3] OR [S4] OR [S5] OR [S6] OR [S7] | 2,321,778 |
| S9 | TITLE("horizon scan" OR "horizon scanning" OR foresight) OR ABSTRACT("horizon scan" OR "horizon scanning" OR foresight) | 5,042 |
| S10 | (TITLE(horizon NEAR/3 scan*) OR ABSTRACT(horizon NEAR/3 scan*)) | 235 |
| S11 | TITLE("foresight practice" OR "foresight method*" OR "strategic foresight") OR ABSTRACT("foresight practice" OR "foresight method*" OR "strategic foresight") | 402 |
| S12 | [S9] OR [S10] OR [S11] | 5,125 |
| S13 | [S12] AND TITLE(search* or scan* or sources or foresight) OR ABSTRACT(search* or scan* or sources or foresight) | 5,125 |
| S14 | [S8] AND [S13] | 576 |

## Appendix 5 WHO’s Global Index Medicus

Databases available: LILACS (Americas) | IMSEAR (South-East Asia) | IMEMR (Eastern Mediterranean) | WPRO (Western Pacific) | AIM (African)

| Searches | Results |
| --- | --- |
| (ti:(equit* OR inequit* OR inequalit* OR disparit* OR equality) OR ab:(equit* OR inequit* OR inequalit* OR disparit* OR equality) OR ti:(ethnic* OR race OR racial* OR racis*) OR ab:(ethnic* OR race OR racial* OR racis*) OR ti:("progress plus" OR ab:"progress plus") OR ti:(ses OR sep OR sociodemographic* OR socio-demographic* OR income OR wealth* OR poverty OR educational level OR level of education OR educational attainment OR well educated OR better educated OR unemploy* OR home owner* OR tenure OR affluen* OR well off OR better off OR worse off OR ab:ses OR sep OR sociodemographic* OR socio-demographic* OR income OR wealth* OR poverty OR educational level OR level of education OR educational attainment OR well educated OR better educated OR unemploy* OR home owner* OR tenure OR affluen* OR well off OR better off OR worse off) OR mh:("Health Inequities" OR "Health Inequities" OR "Gender Equity" OR "Health Status Disparities" OR "Healthcare Disparities" OR "Socioeconomic Disparities in Health" OR "Right to Health" OR "Vulnerable Populations" OR "Health Equity" OR "Urban Population" OR "Poverty" OR "Socioeconomic Factors" OR "Poverty Areas" OR "Social Class" OR "Diversity, Equity, Inclusion")) AND (ti:("horizon scan" OR "horizon scanning" OR foresight) OR ab:("horizon scan" OR "horizon scanning" OR foresight) OR ti:("foresight practice" OR "foresight method*" OR "strategic foresight") OR ab:("foresight practice" OR "foresight method*" OR "strategic foresight") OR mh:("Forecasting")) | 73 |

## Appendix 6 Table of excluded studies

Appendix 6: Table of excluded studies
| Study ID | Title | Reason for exclusion |
| --- | --- | --- |
| Baguant 2019[24] | Integration of Information and Communication Technologies in Teaching by Female Academic Teaching Staff in the Higher Education Sector in Mauritius | Wrong population |
| Daniels et al., 2021[25] | Using foresight to explore the impacts of flooding in Houston on health, poverty, and equity out to 2050 | No framework |
| Fung et al., 2010[26] | Identification of innovation in public health | No framework |
| Graboyes et al., 2020[27] | COVID-19 pandemic and health care disparities in head and neck cancer: Scanning the horizon | Wrong outcome |
| Hemmat et al., 2019[28] | Health information technologies in Iran: Opportunities for development | No framework |
| Hilderink and Eisinger 2021[29] | Dutch Public health Foresight Study, in the light of COVID-19 | Full text not available |
| Instone et al., 2022[30] | International Horizon Scanning the impact of Covid-19 on increasing the health gap and vulnerability | Wrong outcome |
| Jackson et al., 2024[31] | A consensus statement on perinatal mental health during the COVID-19 pandemic and recommendations for post-pandemic recovery and re-build | Wrong outcome |
| Park et al., 2018[32] | The Stakeholder Involvement Strategy For Horizon Scanning In Korea | Full text not available |
| Santana et al., 2020[33] | Advancing tools to promote health equity across European Union regions: the EURO-HEALTHY project | Wrong population |
| Seed 2021[34] | Horizon Scanning in Cancer Genomics: How Advances in Genomic Medicine Will Change Cancer Care Over the Next Decade | No framework |
| Thorpe et al., 2022[35] | Building Public Health Surveillance 3.0: Emerging Timely Measures of Physical, Economic, and Social Environmental Conditions Affecting Health | Wrong outcome |
| Urman et al., 2018[36] | Harnessing AI for health equity in oncology research and practice | Wrong outcome |

## Notes

### Competing Interest Statement

The authors have declared no competing interest.

